# Comparison of surgical quality and long-term outcome between hybrid trans-anal total meso-rectal excision and laparoscopic total meso-rectal excision: a systematic review and meta-analysis

**DOI:** 10.1101/2020.04.29.20085829

**Authors:** Yingchi Yang, Huihui Wang, Kaixin Zhao, Xiangyu Chu, Kai Pang, Yun Yang, Jun Li, Hongwei Yao, Lan Jin, Zhongtao Zhang

## Abstract

**BACKGROUND:** Laparoscopy-assisted trans-anal TME (ta-TME), or hybrid ta-TME, inherited the advantages of both trans-anal surgery and trans-abdominal surgery, and is gaining increasing acceptance from colorectal surgeons worldwide. This research aims to make a comprehensive comparison between hybrid ta-TME surgery and traditional laparoscopic TME (la-TME) surgery regarding surgical quality and long-term survival.

**METHODS:** Cochrane Library, EMbase, Web of Science and PubMed were searched for studies comparing hybrid ta-TME with traditional la-TME. Indicators for surgical quality and long-term prognosis were extracted and pooled. Heterogeneity was assessed with I^2^ index and was significant when p < 0.1 and I^2^ > 50%. Publication bias was estimated by Egger’s test, where p<0.1 was considered statistically significant.

**RESULTS:** 13 studies with 992 patients were included in meta-analysis, of which 467 were in hybrid ta-TME cohorts, and 525 were in traditional la-TME cohorts. Compared with traditional la-TME, hybrid ta-TME has lower rate of positive circumferential margin (RR=0.454, 95%CI 0.240~0.862, p=0.016) and lower conversion rate (RR=0.336, 95%CI 0.134~0.844, p=0.020). On rate of positive distal resection margin, completeness/near-completeness of meso-rectum, overall complications, anal leakage, ileus, urinary dysfunction, 2-year DFS and 2-year OS, there were no significant difference between the two techniques.

**CONCLUSIONS:** Hybrid ta-TME is significantly superior to traditional la-TME in ensuring CRM safety and lowering intra-operative conversion rate, and is meanwhile not inferior on other major outcome indicators concerning surgical quality and long-term survival. To further understand this new surgical technique, we need high-quality RCTs, as well as previous researchers’ updates with results of prolonged follow-up.

## 1. Introduction

Total meso-rectal excision (TME) is the golden standard for the surgical treatment of middle and distal rectal cancer [1]. In 2016, NCCN Practice Guidelines in Oncology: Rectal Cancer made conditional recommendations on the application of laparoscopic surgeries [2], balancing the need for both radical resection and micro-invasiveness. However, for patients with obesity, extra-large tumors, narrow pelvis, male gender or history of neoadjuvant radiotherapy, laparoscopic TME (la-TME) can be particularly difficult to operate, thus undermining the quality of resected specimen. Whereas TME specimen with incomplete meso-rectum, positive circumferential resection margin (CRM), narrow distal resection margin (DRM) or ruptured intestinal wall will naturally increase the risk of local recurrence [3].

With the evolution of concepts and techniques, existing issues in the traditional laparoscopic approach for rectal cancer resection, particularly concerns over the quality of specimen [4], are currently raising increasing awareness. In 2010, Sylla and Atallah respectively reported a radically resected case that were operated trans-anally with the assistance of laparoscopy [5–6], both of which were successful, bringing forward the notion of trans-anal total meso-rectal resection (ta-TME). In pure-NOTES hybrid ta-TME, the operator will have to dissociate around the tumor before prospecting the abdominal cavity, which clearly violated the basic principle for resecting malignant tumors and was thus limited in its application [7]. Hybrid ta-TME, or laparoscopy-assisted hybrid ta-TME, inherited the advantages of both trans-anal surgery and trans-abdominal surgery, and is gaining increasing acceptance from colorectal surgeons worldwide.

To make a comprehensive comparison of surgical quality and long-term survival between traditional laparoscopic TME (la-TME) surgery and hybrid (laparoscopy-assisted) trans-anal TME surgery, key indicators were extracted from relevant studies and pooled for further analysis.

## 2. Methods

### 2.1. Literature search strategy

Protocol of this research was registered with PROSPERO (CRD42020169185). Cochrane Library, EMbase, Web of Science and PubMed were searched for relevant studies published during 2010.1 ~2020.1. Search strategy was adopted as: RECTAL and CANCER or NEOPLASM and TRANSANAL and LAPAROSCOPIC and TOTAL MESORECTAL EXCISION or TME or TA-TME. Each of the literature screening processes was conducted by two researchers, where disputes were settled by consultation with a third senior researcher.

### 2.2. Inclusion and exclusion criteria

Inclusion criteria: studies focusing on rectal cancer patients receiving surgical treatment; with two comparing cohorts (i.e. patients receiving hybrid ta-TME surgeries, and patients receiving la-TME surgeries); language was restricted to English only.

Exclusion criteria: ta-TME cohort contains patients receiving pure-NOTES ta-TME surgeries (i.e. without trans-abdominal procedure at all); ambiguous about whether or not its ta-TME cohort included patients receiving pure-NOTES ta-TME surgeries.

Additionally, studies where ta-TME cohort has excessively long duration of surgery (over 300 minutes, mean or median) were also excluded, as insufficient surgical skillfulness of the new technique might confound the pooled result and bring unnecessary heterogeneity.

### 2.3. Quality evaluation and data extraction

Articles included in this research were all non-randomized controlled trials (non-RCTs). Therefore Newcastle Ottawa Scale (NOS) was adopted for quality evaluation [8].

Items collected from literatures include: author; year of publication; gender ratio; age; BMI; neo-adjuvant therapy (Y/N); ratio of LAR (low anterior resection) procedure; duration of surgery; incidence of CRM+, DRM+, completeness/near-completeness of mesorectum, overall complications, anal leakage, ileus, urinary dysfunction, conversion to open surgery, re-operation, 2-year disease free survival (DFS) and 2-year overall survival (OS).

### 2.4. Statistics

Statistical analyses in this research included pooling of outcomes, test of heterogeneity and assessment of publication bias. Pooled analysis was conducted for each selected outcome indicator. All indicators were dichotomous data, therefore relative risk with corresponding 95% confidence interval were calculated. Heterogeneity was assessed with I^2^ index, where p < 0.1 and I^2^ > 50% was considered significant and a random-effect model (Inverse Variance) was thus adopted; otherwise a fixed-effect model (Mantel-Haenszel) was used [9, 10]. Exclusion of a study due to significant heterogeneity was forbidden, so as to display a comprehensive analysis result that was true to the study screening mechanism. Publication bias was estimated by Egger’s test, and was considered significant when p<0.1 [11, 12]. All analysis was processed on STATA software, version 15.0 (StataCorp LP, College Station, TX, USA). Initial results were further validated with MedCalc, version 19.1.3 (MedCalc Software, Ostend, Belgium).

## 3. Results

### 3.1. Search results

13 cohort studies [13–25] were included based on the above selection mechanism. 9 out of the 13 were matched case-control studies (propensity score matching on a 1:1 basis). Literature selection process is demonstrated in Figure 1. Characteristics of included studies and patients were displayed in Table 1.

**Fig 1.**
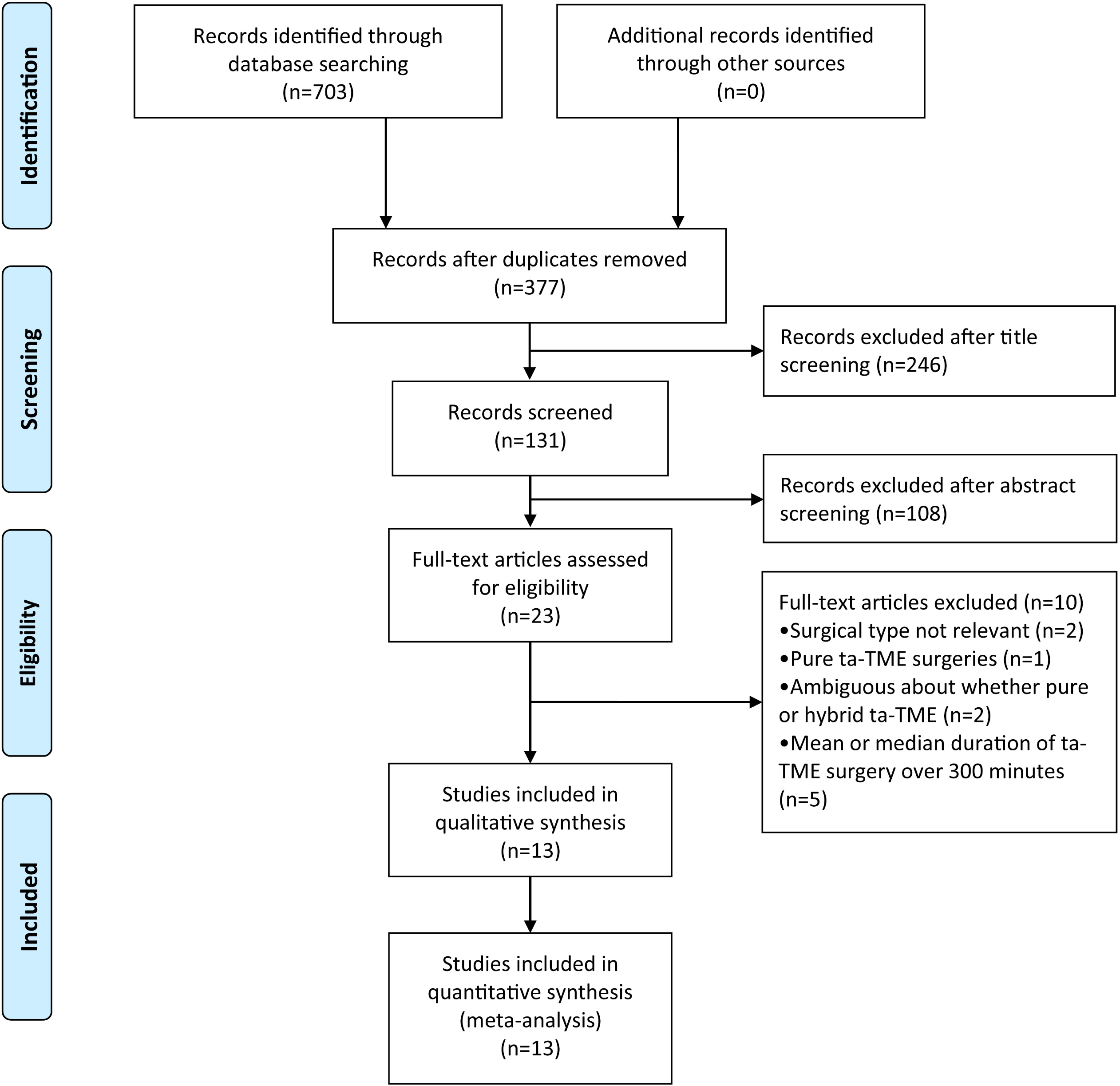
Flowchart of the literature screening process.

**Table 1.**
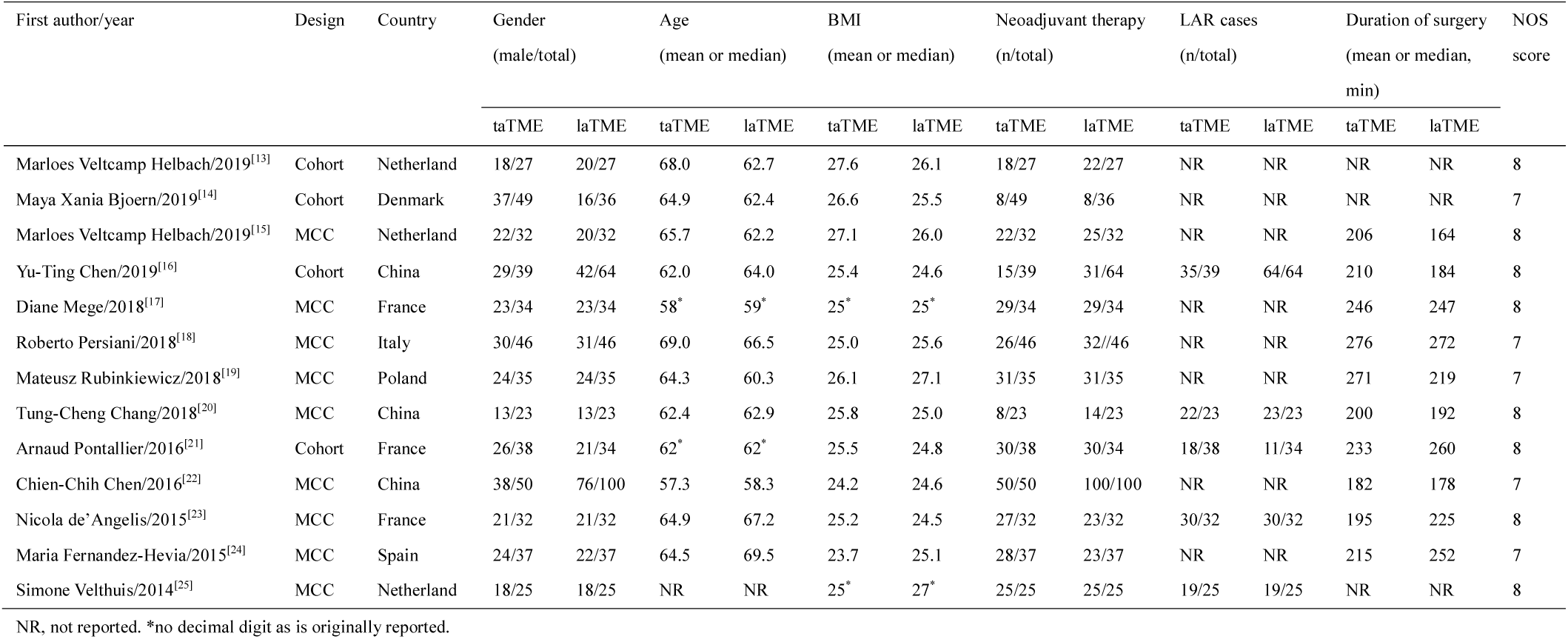
Characteristics of included studies

| First author/year | Design | Country | Gender |  | Age |  | BMI |  | Neoadjuvant therapy |  | LAR cases |  | Duration of surgery |  | NOS score |
| --- | --- | --- | --- | --- | --- | --- | --- | --- | --- | --- | --- | --- | --- | --- | --- |
|  |  |  | (male/total) |  | (mean or median) |  | (mean or median) |  | (n/total) |  | (n/total) |  | (mean or median, min) |  |  |
|  |  |  | taTME | laTME | taTME | laTME | taTME | laTME | taTME | laTME | taTME | laTME | taTME | laTME |  |
| Marloes Veltcamp Helbach/2019<sup>[13]</sup> | Cohort | Netherland | 18/27 | 20/27 | 68.0 | 62.7 | 27.6 | 26.1 | 18/27 | 22/27 | NR | NR | NR | NR | 8 |
| Maya Xania Bjoern/2019<sup>[14]</sup> | Cohort | Denmark | 37/49 | 16/36 | 64.9 | 62.4 | 26.6 | 25.5 | 8/49 | 8/36 | NR | NR | NR | NR | 7 |
| Marloes Veltcamp Helbach/2019<sup>[15]</sup> | MCC | Netherland | 22/32 | 20/32 | 65.7 | 62.2 | 27.1 | 26.0 | 22/32 | 25/32 | NR | NR | 206 | 164 | 8 |
| Yu-Ting Chen/2019<sup>[16]</sup> | Cohort | China | 29/39 | 42/64 | 62.0 | 64.0 | 25.4 | 24.6 | 15/39 | 31/64 | 35/39 | 64/64 | 210 | 184 | 8 |
| Diane Mege/2018<sup>[17]</sup> | MCC | France | 23/34 | 23/34 | 58* | 59* | 25* | 25* | 29/34 | 29/34 | NR | NR | 246 | 247 | 8 |
| Roberto Persiani/2018<sup>[18]</sup> | MCC | Italy | 30/46 | 31/46 | 69.0 | 66.5 | 25.0 | 25.6 | 26/46 | 32//46 | NR | NR | 276 | 272 | 7 |
| Mateusz Rubinkiewicz/2018<sup>[19]</sup> | MCC | Poland | 24/35 | 24/35 | 64.3 | 60.3 | 26.1 | 27.1 | 31/35 | 31/35 | NR | NR | 271 | 219 | 7 |
| Tung-Cheng Chang/2018<sup>[20]</sup> | MCC | China | 13/23 | 13/23 | 62.4 | 62.9 | 25.8 | 25.0 | 8/23 | 14/23 | 22/23 | 23/23 | 200 | 192 | 8 |
| Arnaud Pontallier/2016<sup>[21]</sup> | Cohort | France | 26/38 | 21/34 | 62* | 62* | 25.5 | 24.8 | 30/38 | 30/34 | 18/38 | 11/34 | 233 | 260 | 8 |
| Chien-Chih Chen/2016<sup>[22]</sup> | MCC | China | 38/50 | 76/100 | 57.3 | 58.3 | 24.2 | 24.6 | 50/50 | 100/100 | NR | NR | 182 | 178 | 7 |
| Nicola de' Angelis/2015<sup>[23]</sup> | MCC | France | 21/32 | 21/32 | 64.9 | 67.2 | 25.2 | 24.5 | 27/32 | 23/32 | 30/32 | 30/32 | 195 | 225 | 8 |
| Maria Fernandez-Hevia/2015<sup>[24]</sup> | MCC | Spain | 24/37 | 22/37 | 64.5 | 69.5 | 23.7 | 25.1 | 28/37 | 23/37 | NR | NR | 215 | 252 | 7 |
| Simone Velthuis/2014<sup>[25]</sup> | MCC | Netherland | 18/25 | 18/25 | NR | NR | 25* | 27* | 25/25 | 25/25 | 19/25 | 19/25 | NR | NR | 8 |

NR, not reported. \*no decimal digit as is originally reported.

### 3.2. Meta-analysis result

#### 3.2.1. Quality of resected specimen

##### CRM+

12 studies compared the rate of CRM+ between hybrid ta-TME and traditional laparoscopic TME. Pooled rate of CRM+ of hybrid ta-TME is significantly lower than that of traditional laparoscopic TME (Relative Risk=0.454, 95%CI 0.240~0.862, p=0.016) (Figure 2). Analysis was conducted with a fixed-effect model as no significant heterogeneity was observed (p=0.849, I^2^= 0.0%). Egger’s test for publication bias is negative (p=0.509).

**Fig 2.**
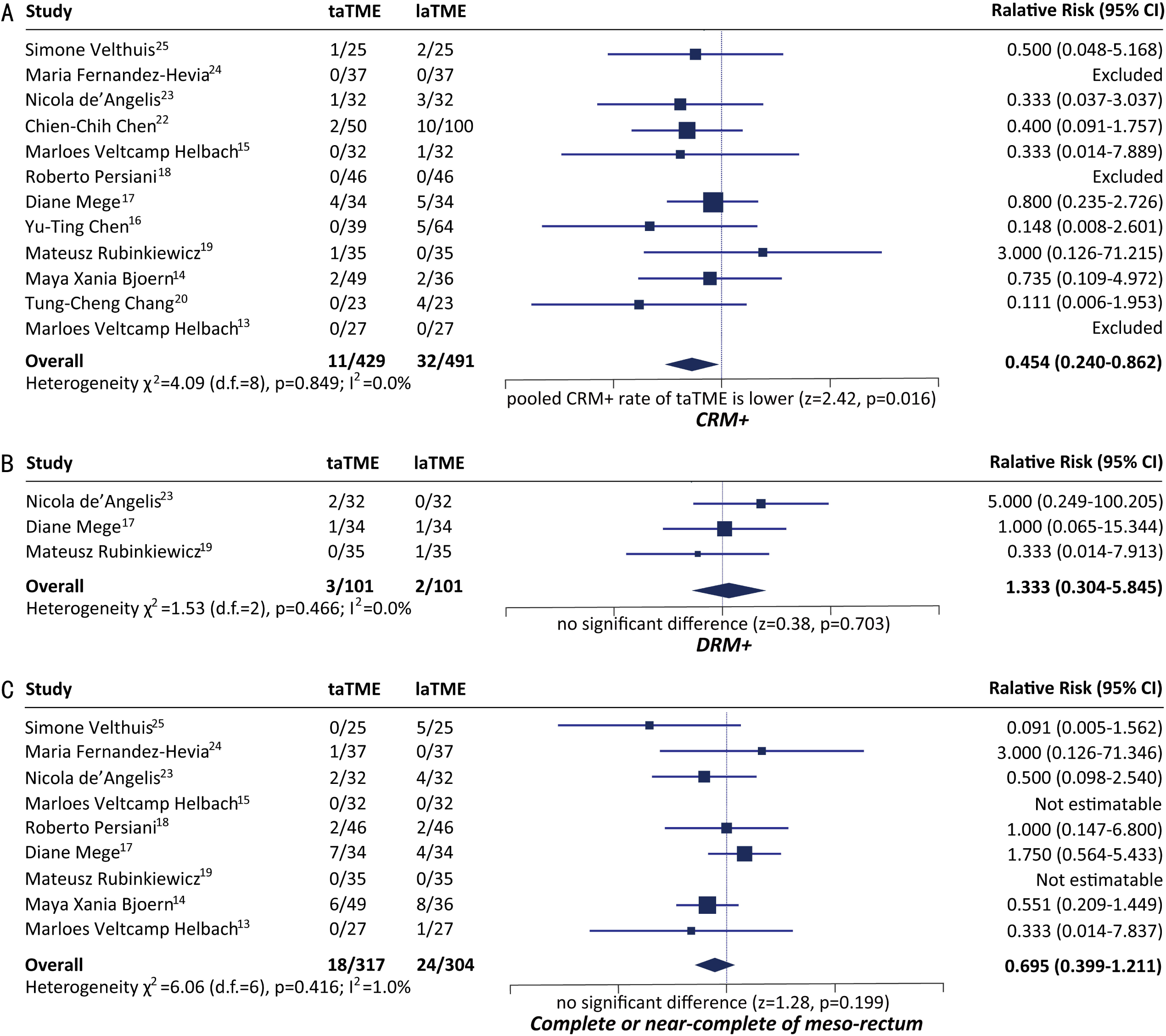
Comparison of quality of resected specimen.

##### DRM+

3 studies compared the rate of DRM+ between hybrid ta-TME and traditional laparoscopic TME, and pooled result indicated no significant difference between the two techniques (Relative Risk=1.333, 95%CI 0.304~5.845, p=0.703) (Figure 2). Analysis was conducted with a fixed-effect model as no significant heterogeneity was observed (p=0.466, I^2^ = 0.0%). Egger’s test was infeasible due to insufficiency of studies.

##### Integrity of mesorectum

9 studies compared the complete/near-complete rate of meso-rectum between hybrid ta-TME and traditional laparoscopic TME, and pooled result indicated no significant difference between the two techniques (Relative Risk=0.695, 95%CI 0.399~1.211, p=0.199) (Figure 2). Analysis was conducted with a fixed-effect model as no significant heterogeneity was observed (p=0.416, I^2^= 1.0%). Egger’s test for publication bias is negative (p=0.934).

#### 3.2.2. Post-operative complications

##### Overall complications

11 studies compared the incidence rate of overall post-operative complications between hybrid ta-TME and traditional laparoscopic TME, and pooled result indicated no significant difference between the two techniques (Relative Risk=0.784, 95%CI 0.612~1.004, p=0.054) (Figure 3). Analysis was conducted with a fixed-effect model as no significant heterogeneity was observed (p=0.943, I^2^= 0.0%). Egger’s test for publication bias is negative (p=0.510).

**Fig 3.**
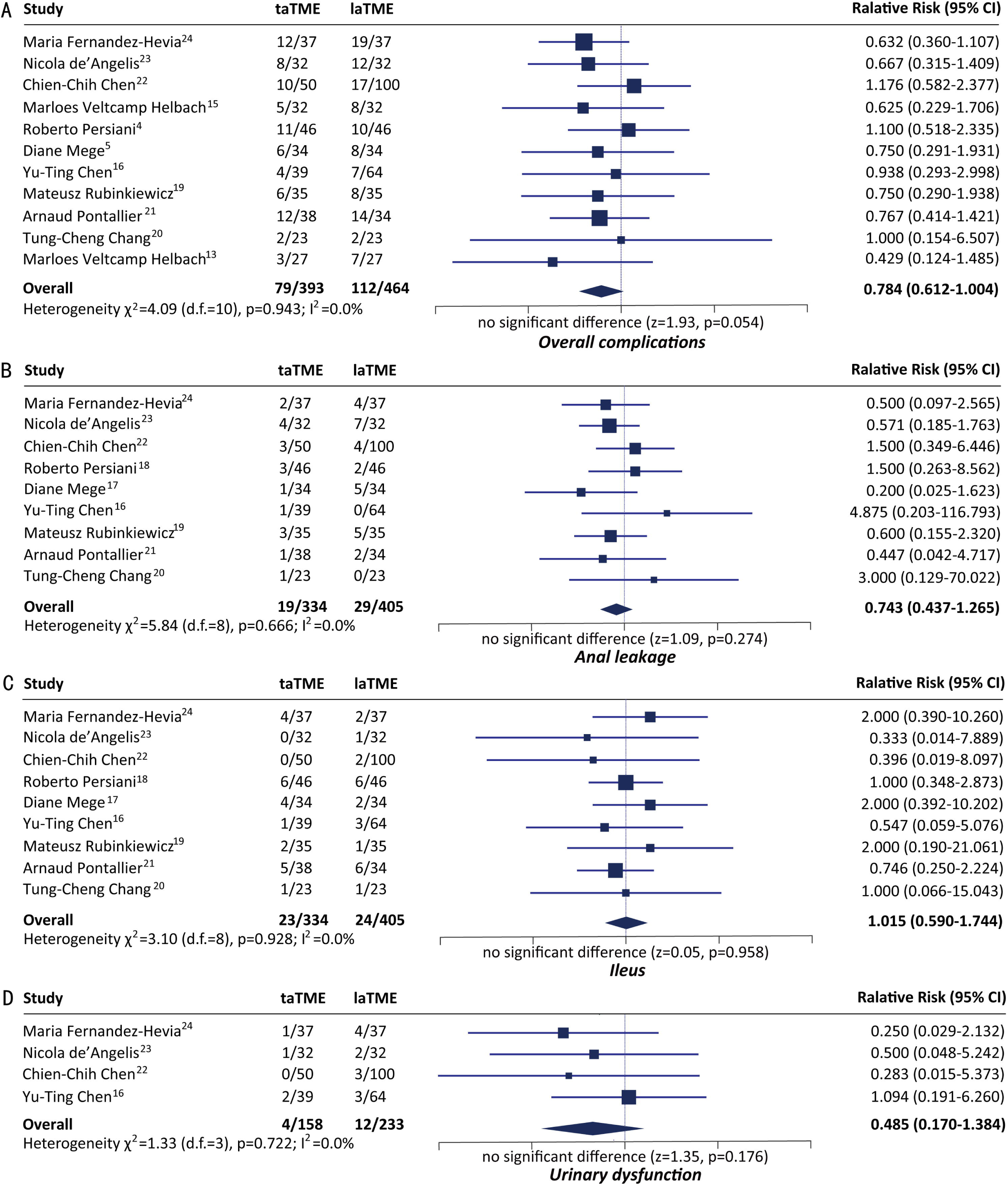
Comparison of incidence of post-operative complications.

##### Anal leakage

9 studies compared the incidence rate of post-operative anal leakage between hybrid ta-TME and traditional laparoscopic TME, and pooled result indicated no significant difference between the two techniques (Relative Risk=0.743, 95%CI 0.437~1.265, p=0.274) (Figure 3). Analysis was conducted with a fixed-effect model as no significant heterogeneity was observed (p=0.666, I^2^ =0.0%). Egger’s test for publication bias is negative (p=0.485).

##### Ileus

9 studies compared the incidence rate of post-operative ileus between hybrid ta-TME and traditional laparoscopic TME, and pooled result indicated no significant difference between the two techniques (Relative Risk=1.015, 95%CI 0.590~1.744, p=0.958) (Figure 3). Analysis was conducted with a fixed-effect model as no significant heterogeneity was observed (p=0.928, I^2^ =0.0%). Egger’s test for publication bias is negative (p=0.556).

##### Urinary dysfunction

4 studies compared the incidence rate of post-operative urinary dysfunction between hybrid ta-TME and traditional laparoscopic TME, and pooled result indicated no significant difference between the two techniques (Relative Risk=0.485, 95%CI 0.170~1.384, p=0.176) (Figure 3). Analysis was conducted with a fixed-effect model as no significant heterogeneity was observed (p=0.722, I^2^ =0.0%). Egger’s test for publication bias is negative (p=0.463).

#### 3.2.3. Unplanned surgical events

Conversion to open surgery

7 studies compared the conversion rate between hybrid ta-TME and traditional laparoscopic TME. Pooled conversion rate of hybrid ta-TME is significantly lower than that of traditional laparoscopic TME (Relative Risk=0.336, 95%CI 0.134~0.844, p=0.020) (Figure 4). Analysis was conducted with a fixed-effect model as no significant heterogeneity was observed (p=0.404, I^2^=0.4%). Egger’s test for publication bias is negative (p=0.243).

**Fig 4.**
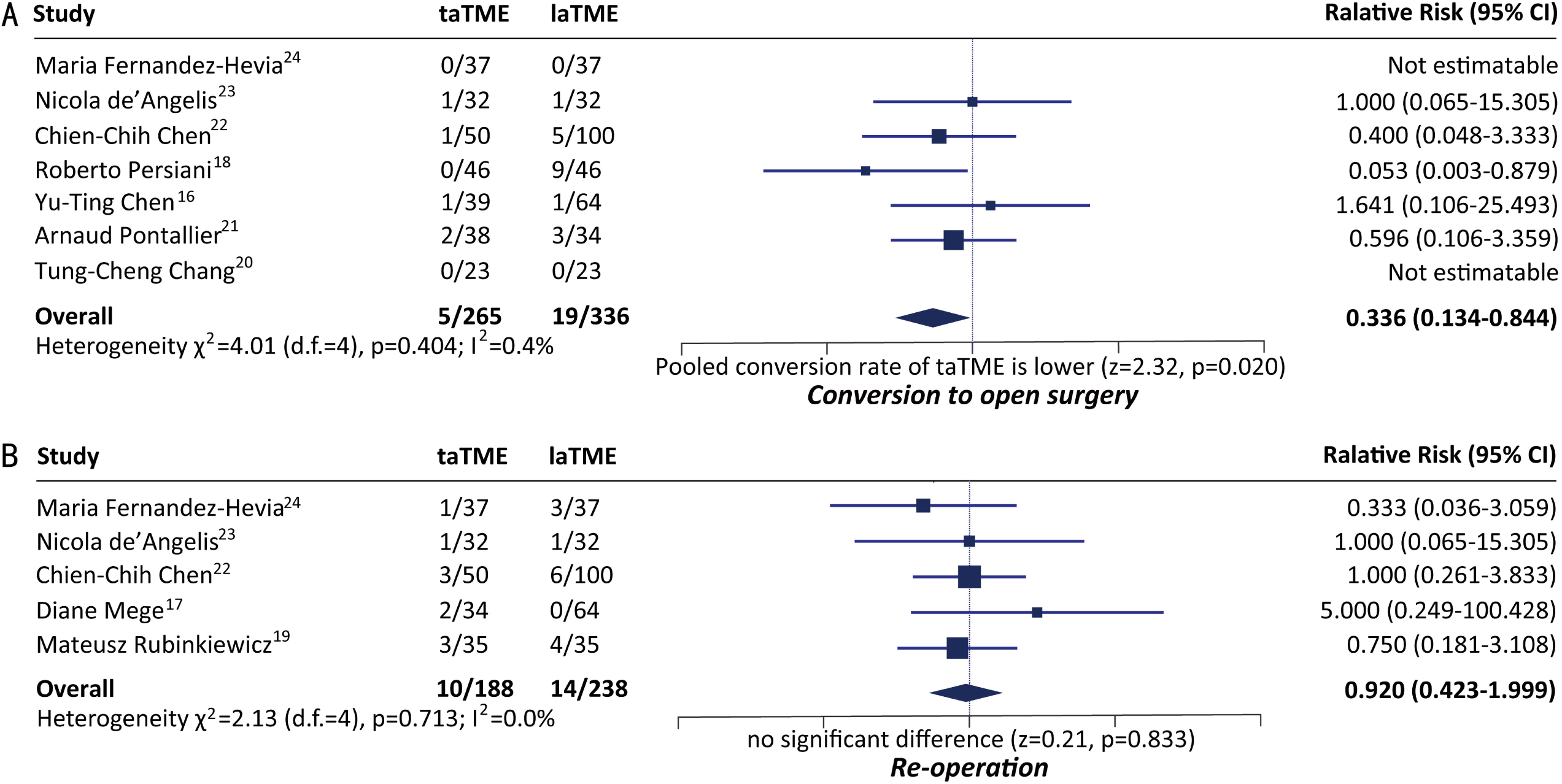
Comparison of incidence of unplanned surgical events.

##### Re-operation

5 studies compared the incidence rate of unplanned re-operation between hybrid ta-TME and traditional laparoscopic TME, and pooled result indicated no significant difference between the two techniques (Relative Risk=0.920, 95%CI 0.423~1.999, p=0.833) (Figure 4). Analysis was conducted with a fixed-effect model as no significant heterogeneity was observed (p=0.713, I^2^=0.0%). Egger’s test for publication bias is negative (p=0.577).

#### 3.2.4. Long-term oncological outcome

Two-year DFS.

2 studies compared the two-year DFS between hybrid ta-TME and traditional laparoscopic TME, and pooled result indicated no significant difference between the two techniques (Relative Risk=1.017, 95%CI 0.914~1. 131, p=0.757) (Figure 5). Analysis was conducted with a fixed-effect model as no significant heterogeneity was observed (p=0.530, I^2^ =0.0%). Egger’s test was infeasible due to insufficiency of studies.

**Fig 5.**
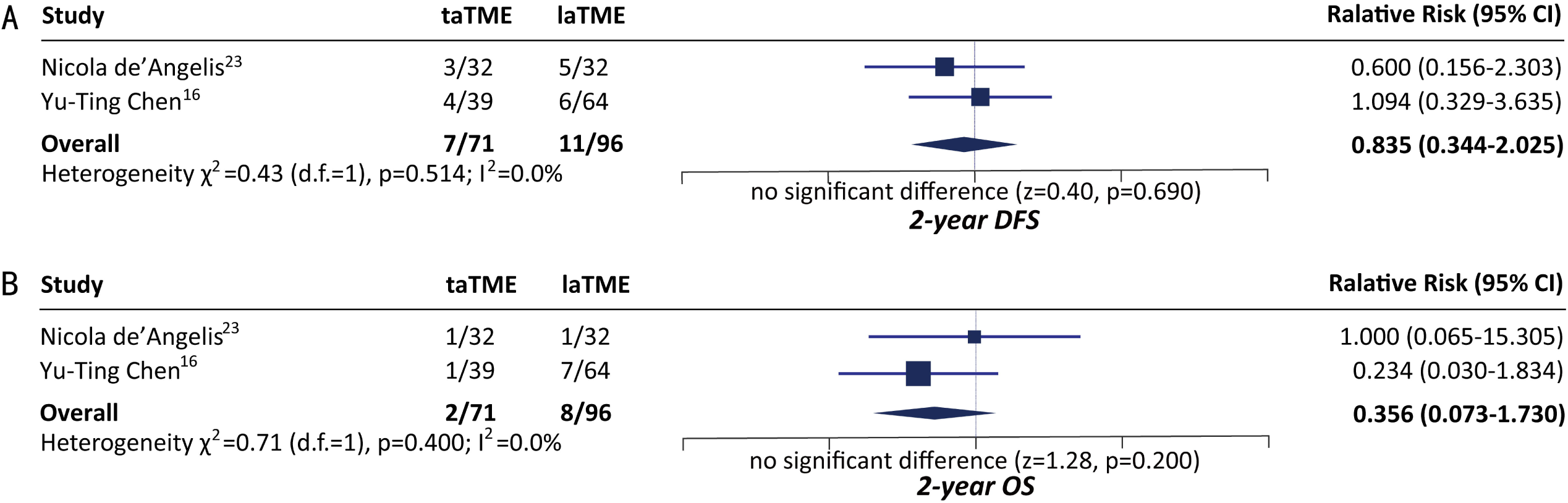
Comparison of long-term survival.

##### Two-year OS

2 studies compared two-year OS between hybrid ta-TME and traditional laparoscopic TME, and pooled result indicated no significant difference between the two techniques (Relative Risk=1.048, 95%CI 0.974~1.127, p=0.211) (Figure 5). Analysis was conducted with a fixed-effect model as no significant heterogeneity was observed (p=0.170, I^2^ =46.8%). Egger’s test was infeasible due to insufficiency of studies.

## 4. Discussion

As indicated by the results of meta-analysis, pooled CRM+ rate of hybrid ta-TME is significantly lower than that of la-TME, which is in accordance with our empirical understanding that ta-TME better ensures the safety of circumferential resection margin. However, no significant differences were observed on rate of DRM+, as well as rate of completeness/near-completeness of mesorectum. Regarding overall post-operative complications, post-operative ileus, post-operative anal leakage and post-operative urinary dysfunction, there are no significant differences between hybrid ta-TME and traditional la-TME. Yet it’s worth mentioning that among these four indicators, the difference on rate of overall complications is close to reaching significance (p=0.054). As to unplanned surgical events, the pooled conversion rate (ratio of cases converted to open surgery) of hybrid ta-TME is significantly lower than that of la-TME, indicating ta-TME’s distinct advantages in handling complex surgical scenarios over la-TME. Nonetheless, no significant difference was observed on rate of re-operation, suggesting that there was no significant difference between hybrid ta-TME and traditional la-TME on severe post-operative complications. Last but not the least, as the primary concern of clinical physicians, difference on 2-year DFS and 2-year OS between the two surgical strategies were insignificant. Notably, significant heterogeneity and publication bias were observed in pooling of none of the above indicators, indicating good credibility of the results (Table 2).

**Table 2.** A summary of pooled analysis results

| Pooled indicators | Overall significance |  | Heterogeneity |  |  | Publication bias |  |
| --- | --- | --- | --- | --- | --- | --- | --- |
| | p value of test of RR | Significant or not<sup>#</sup> (Y/N) | p value of heterogeneity $\chi^2$ | I<sup>2</sup> | Significant or not<sup>#</sup> (Y/N) | p value of Egger's test | Significant or not<sup>#</sup> (Y/N) |
| CRM+ | 0.016 | Y | 0.849 | 0.0% | N | 0.509 | N |
| DRM+ | 0.703 | N | 0.466 | 0.0% | N | NA* | NA* |
| Complete/Near-complete of meso-rectum | 0.199 | N | 0.171 | 33.6% | N | 0.934 | N |
| Overall complications | 0.054 | N | 0.943 | 0.0% | N | 0.510 | N |
| Anal leakage | 0.274 | N | 0.666 | 0.0% | N | 0.485 | N |
| Ileus | 0.958 | N | 0.928 | 0.0% | N | 0.556 | N |
| Urinary dysfunction | 0.176 | N | 0.722 | 0.0% | N | 0.463 | N |
| Conversion to open surgery | 0.020 | Y | 0.404 | 0.4% | N | 0.243 | N |
| Re-operation | 0.833 | N | 0.713 | 0.0% | N | 0.577 | N |
| 2-year DFS | 0.690 | N | 0.530 | 0.0% | N | NA* | NA* |
| 2-year OS | 0.200 | N | 0.170 | 46.8% | N | NA* | NA* |

CRM circumferential resection margin, DRM distal resection margin, DFS disease free survival, OS overall survival, Y yes, N no, NA not applicable. \* Egger's test not conducted due to insufficiency of studies, # Test of RR was significant when $p < 0.05$ ; Test of heterogeneity was significant when $p < 0.1$ and $I^2 > 50\%$ ; Egger's test was significant when $p < 0.1$ .

Quality of TME surgeries depends heavily on the integrity of resected mesorectum [26], and rectal surgery’s higher demand on intra-operative visual field and quality of specimen prompted the rapid development of ta-TME [27]. The trans-anal procedure started directly from the otherwise most difficult part of TME surgeries, providing clearer field of view and broader space to maneuver. Operating trans-anally can ensure sufficient length of distal margin and guarantee the effectiveness of the radical resection, meanwhile increase the anus-preserving rate of patients with low or ultra-low rectal cancers. This down-to-up strategy allows, even on male patients with narrow pelvis, for easy dissociation along the Denonvilliers fascia after cutting open the rectal wall, decreasing the risks of injuring prostate gland, seminal vesicle, posterior wall of vagina, and pelvic plexus. Hybrid ta-TME, as the combination of trans-abdominal surgery and trans-anal surgery, inherited the advantages of both. In a regular hybrid ta-TME surgery, laparoscopic team prospects the abdominal cavity, ligatures blood vessels, dissect lymph nodes and dissociate the proximal and middle mesorectum; and the trans-anal team dissociate distal mesorectum and retract specimen via natural orifice.

Compared with traditional laparoscopic TME surgeries, hybrid ta-TME significantly decreased the difficulty in a series of surgical actions, including exposing in pelvic bottom, ensuring completeness of resected meso-rectum, anticipating distal resection margin and severing rectum [28–30]. From an empirical point of view, we believe hybrid ta-TME is advantageous over traditional laparoscopic TME primarily in 5 aspects: (1) can accurately sever distal rectum from within rectal cavity, better ensuring the safety of distal resection margin; (2) enters the interstices around distal meso-rectum more conveniently, increasing the safety of circumferential resection margin; (3) does not require an extra incision on the abdomen for specimen retraction, contributing to the idea of micro-invasiveness; (4) reduces usage of anastomosis devices and alleviate patients’ costs; (5) avoids multiple triggering of linear stapling devices, potentially lowering risk of anal leakage [31–33]. The advantages described in (3) and (4) were obvious and need no further explanations. And as results of our meta-analysis indicated, pooled rate of CRM+ of hybrid ta-TME is significantly lower than that of traditional laparoscopic TME (Relative Risk=0.454, 95%CI 0.240~0.862, p=0.016), supporting our empirical statement in (2). However, statements in (1) and (5) were not supported by the pooled results, which may need higher-level evidences from further research.

Nevertheless, hybrid ta-TME surgery also has apparent flaws: (1) trans-anal procedures are time-consuming, and requires abundant experiences of single-port laparoscopic surgeries, dictating a prolonged learning curve; (2) operating trans-anally can obviously increase the risks of pelvic and abdominal infection; (3) when operating trans-anally, judgement on the location of the end of meso-rectum is very difficult to make [34], which may cause, precisely opposite to the original purpose of ta-TME, incomplete resection of meso-rectum. Naturally, towards a new technique, there will be constant doubts and questioning. However, there will never be a perfect technique that befit all patients. Only by actively selecting the most suitable patients and determining the optimal indications for ta-TME can we maximize its efficacy. Currently, existing studies on ta-TME technique are mostly retrospective studies with small-sized samples. In 2015, an international, multi-centered, prospective, random controlled trial code-named COLOR was initiated by researchers from Netherland. COLOR aimed to include 1098 mid and lower rectal cancer patients from around the globe to compare the safety and efficacy of ta-TME and traditional laparoscopic TME [35]. As a participating center of COLOR, our hospital founded the *Steering Committee of COLOR Trial in China* so as to provide appropriate assistance to interested centers across China. Meanwhile, we had established the national ta-TME database of China, which has now registered over 1000 ta-TME cases from 40 domestic centers, hoping to provide more higher-level evidence for the development of ta-TME technique.

In conclusion, hybrid ta-TME is significantly superior to traditional la-TME in ensuring CRM safety and lowering intra-operative conversion rate. But as to the rate of DRM+, completeness/near-completeness of meso-rectum, overall complications, anal leakage, ileus, urinary dysfunction, 2-year DFS and 2-year OS, there are no significant difference between the two TME techniques. To further understand this novel surgical strategy, we need high-quality RCTs, as well as previous researchers’ updates with results of prolonged follow-up.

## Data Availability

All research data are available via reasonable request by email to corresponding author.

## Referrences

1. Heald RJ, Husband EM, Ryall RD. The mesorectum in rectal cancer surgery—the clue to pelvic recurrence? Br J Surg. 1982 Oct;69(10):613–6.

2. National Comprehensive Cancer Network. NCCN Practice Guidelines in Oncology: rectal cancer. Version.1. Washington: National Comprehensive Cancer Network; 2016.

3. Bosch SL, Nagtegaal ID. The importance of the pathologist’ s role in assessment of the quality of the mesorectum. Curr Colorectal Cancer Rep. 2012 Jun;8(2):90–8.

4. Fleshman J, Branda M, Sargent DJ, Boller AM, George V, Abbas M, et al. Effect of laparoscopicassisted resection vs open resection of stage II or III rectal cancer on pathologic outcomes: the ACOSOG Z6051 randomized clinical trial. JAMA. 2015 Oct;314(13):1346—55.

5. Sylla P, Rattner DW, Delgado S, Lacy AM. NOTES transanal rectal cancer resection using transanal endoscopic microsurgery and laparoscopic assistance. Surg Endosc. 2010 May;24(5):1205–10.

6. Atallah S, Albert M, Larach S. Transanal minimally invasive surgery: a giant leap forward. Surg Endosc. 2010 Sep;24(9):2200–5.

7. Marks J, Mizrahi B, Dalane S, Nweze I, Marks G. Laparoscopic transanal abdominal transanal resection with sphincter preservation for rectal cancer in the distal 3 cm of the rectum after neoadjuvant therapy. Surg Endosc. 2010 Nov;24(11):2700–7.

8. Stang A. Critical evaluation of the Newcastle-Ottawa scale for the assessment of the quality of nonrandomized studies in meta-analyses. Eur J Epidemiol. 2010 Sep;25(9):603–5.

9. Higgins JP, Green S, editors. Cochrane Handbook for Systematic Reviews of Interventions Version 5.1.0 (updated March 2011). The Cochrane Collaboration, 2011. Available from http://www.cochrane-handbook.org

10. Higgins JP, Thompson SG, Deeks JJ, Altman DG. Measuring inconsistency in meta-analyses. BMJ. 2003 Sep;327(7414):557–60.

11. Hayashino Y, Noguchi Y, Fukui T. Systematic evaluation and comparison of statistical tests for publication bias. J Epidemiol. 2005 Nov;15(6):235–43.

12. Egger M, Davey Smith G, Schneider M, Minder C. Bias in meta-analysis detected by a simple, graphical test. BMJ. 1997 Sep;315(7109):629–34.

13. Veltcamp Helbach M, Koedam TW, Knol JJ, Velthuis S, Bonjer HJ, Tuynman JB, et al. Quality of life after rectal cancer surgery: differences between laparoscopic and transanal total mesorectal excision. Surg Endosc. 2019 Jan;33(1):79–87.

14. Bjoern MX, Nielsen S, Perdawood SK. Quality of Life After Surgery for Rectal Cancer: a Comparison of Functional Outcomes After Transanal and Laparoscopic Approaches. J Gastrointest Surg. 2019 Aug;23(8):1623–30.

15. Veltcamp Helbach M, Koedam TW, Knol JJ, Diederik A, Spaargaren GJ, Bonjer HJ, et al. Residual mesorectum on postoperative magnetic resonance imaging following transanal total mesorectal excision (TaTME) and laparoscopic total mesorectal excision (LapTME) in rectal cancer. Surg Endosc. 2019 Jan;33(1):94–102.

16. Chen YT, Kiu KT, Yen MH, Chang TC. Comparison of the short-term outcomes in lower rectal cancer using three different surgical techniques: transanal total mesorectal excision (TME), laparoscopic TME, and open TME. Asian J Surg. 2019 Jun;42(6):674–80.

17. Mege D, Hain E, Lakkis Z, Maggiori L, Prost À la Denise J, Panis Y. Is trans-anal total mesorectal excision really safe and better than laparoscopic total mesorectal excision with a perineal approach first in patients with low rectal cancer? A learning curve with case-matched study in 68 patients. Colorectal Dis. 2018 Jun;20(6):O143–51.

18. Persiani R, Biondi A, Pennestrì F, Fico V, De Simone V, Tirelli F, et al. Transanal Total Mesorectal Excision vs Laparoscopic Total Mesorectal Excision in the Treatment of Low and Middle Rectal Cancer: A Propensity Score Matching Analysis. Dis Colon Rectum. 2018 Jul;61(7):809–16.

19. Rubinkiewicz M, Nowakowski M, Wierdak M, Mizera M, Dembiński M, Pisarska M, et al. Transanal total mesorectal excision for low rectal cancer: a case-matched study comparing TaTME versus standard laparoscopic TME. Cancer Manag Res. 2018 Nov;10:5239–45.

20. Chang TC, Kiu KT. Transanal Total Mesorectal Excision in Lower Rectal Cancer: Comparison of Short-Term Outcomes with Conventional Laparoscopic Total Mesorectal Excision. J Laparoendosc Adv Surg Tech A. 2018 Apr;28(4):365–9.

21. Pontallier A, Denost Q, Van Geluwe B, Adam JP, Celerier B, Rullier E. Potential sexual function improvement by using transanal mesorectal approach for laparoscopic low rectal cancer excision. Surg Endosc. 2016 Nov;30(11):4924–33.

22. Chen CC, Lai YL, Jiang JK, Chu CH, Huang IP, Chen WS, et al. Transanal Total Mesorectal Excision Versus Laparoscopic Surgery for Rectal Cancer Receiving Neoadjuvant Chemoradiation: A Matched Case-Control Study. Ann Surg Oncol. 2016 Apr;23(4):1169–76.

23. de’Angelis N, Portigliotti L, Azoulay D, Brunetti F. Transanal total mesorectal excision for rectal cancer: a single center experience and systematic review of the literature. Langenbecks Arch Surg. 2015 Dec;400(8):945–59.

24. Fernández-Hevia M, Delgado S, Castells A, Tasende M, Momblan D, Díaz del Gobbo G, et al. Transanal total mesorectal excision in rectal cancer: short-term outcomes in comparison with laparoscopic surgery. Ann Surg. 2015 Feb;261(2):221–7.

25. Velthuis S, Nieuwenhuis DH, Ruijter TE, Cuesta MA, Bonjer HJ, Sietses C. Transanal versus traditional laparoscopic total mesorectal excision for rectal carcinoma. Surg Endosc. 2014 Dec;28(12):3494–9.

26. Nacion AJ, Park YY, Yang SY, Kim NK. Critical and challenging issues in the surgical management of low-lying rectal cancer. Yonsei Med J. 2018 Aug;59(6):703–16.

27. Sylla P, Rattner DW, Delgado S, Lacy AM. NOTES transanal rectal cancer resection using transanal endoscopic microsurgery and laparoscopic assistance. Surg Endosc. 2010 May;24(5):1205–10.

28. de Lacy AM, Rattner DW, Adelsdorfer C, Tasende MM, Fernández M, Delgado S, et al. Transanal natural orifice transluminal endoscopic surgery (NOTES) rectal resection: “down-to-up” total mesorectal excision (TME)—short-term outcomes in the first 20 cases. Surg Endosc. 2013 Sep;27(9):3165–72.

29. Chouillard E, Chahine E, Khoury G, Vinson-Bonnet B, Gumbs A, Azoulay D, et al. NOTES total mesorectal excision (TME) for patients with rectal neoplasia: a preliminary experience. Surg Endosc. 2014 Nov;28(11):3150–7.

30. Wolthuis AM, Bislenghi G, de Buck van Overstraeten A, D’Hoore A. Transanal total mesorectal excision: towards standardization of technique. World J Gastroenterol. 2015 Nov;21(44):12686–95.

31. Park JS, Choi GS, Kim SH, Kim HR, Kim NK, Lee KY, et al. Multicenter analysis of risk factors for anastomotic leakage after laparoscopic rectal cancer excision: the Korean laparoscopic colorectal surgery study group. Ann Surg. 2013 Apr;257(4):665–71.

32. Kawada K, Hasegawa S, Hida K, Hirai K, Okoshi K, Nomura A, et al. Risk factors for anastomotic leakage after laparoscopic low anterior resection with DST anastomosis. Surg Endosc. 2014 Oct;28(10):2988–95.

33. Braunschmid T, Hartig N, Baumann L, Dauser B, Herbst F. Influence of multiple stapler firings used for rectal division on colorectal anastomotic leak rate. Surg Endosc. 2017 Dec;31(12):5318–26.

34. Chi P, Chen Z, Lu X. Transanal total mesorectal excision: can it achieve the standard of TME? Ann Surg. 2017 Dec;266(6):e87–8.

35. Deijen CL, Velthuis S, Tsai A, Mavroveli S, de Lange-de Klerk ES, Sietses C, et al. COLOR III: a multicentre randomised clinical trial comparing transanal TME versus laparoscopic TME for mid and low rectal cancer. Surg Endosc. 2016 Aug;30(8):3210–5.

